## Supplementary material for "Costs, catastrophic out-of-pocket payments and impoverishment related to accessing surgical care among households in rural Ethiopia"

**Supplementary table 1 .Household consumption expenditure cost categories ( in Birr)**

| Expenditure categories | Mean (SD) | Percentage share |
| --- | --- | --- |
| Food | 5970.3(4400.3) | 43.8 |
| Household utilities | 2423.3(7981.7) | 17.7 |
| Big purchase | 1694.9(6830.9) | 12.4 |
| Healthcare(direct medical and non-medical) | 2876.1 (3848.9) | 21.2 |
| Total | 12964.7 (15286.1) |  |

*1 USD = 29.07 Birr (2019, purchasing power parity, 1 international dollars=10.74 Birr)*

*Food items (staple foods, vegetable, fruit, spices etc.), regular household expenses (electricity, water, cooking, renting, clothing, transport, etc.), Big purchase include (education, durable goods, cultural ceremonies, entertainment, tax.); health expenditure include expense on (outpatient consultation, medication, investigations, hospitalization, medical appliances, ambulance, hospital food, accommodation and transport cost related to surgical care)*

Supplementary table 2. Poverty impact of OOP payments for surgical care

| Poverty impact of OOP payments | Half(1/2) of median total consumption | Two -thirds(2/3) of median total consumption |
| --- | --- | --- |
| Poverty head count |  |  |
| Pre-payment head count ^X^ | 18.7% | 29.1% |
| Post payment head count ^Y^ | 29.1% | 48.3% |
| Absolute percentage point change(head count impact) ^Z^_(=Y-X)_ | 10.4% | 19.2% |
| Relative percentage change (=Z/X*100) | 55.6% | 65.9% |
| Poverty gaps |  |  |
| Prepayment poverty gap(Birr) ^X^ | 315.0 | 666.5 |
| Post payment poverty gap(Birr) ^Y^ | 604.9 | 1216.9 |
| Absolute point change(Birr))^Z^_(=Y-X)_ | 289.9 | 550.4 |
| Relative percentage change (=Z/X*100) | 92.0% | 82.5% |
| Normalized poverty gaps |  |  |
| Pre-payment normalized gap ^X^ | 6.6% | 10.4% |
| Mean positive pre-payment poverty gap(Birr) | 1686.4 | 2511.5 |
| Post-payment normalized gap ^Y^ | 12.6% | 19.1 |
| Mean positive post-payment poverty gap(Birr) | 2077.4 | 2516.8 |
| Absolute percentage point change (impact) ^Z^_(=Y-X)_ | 6.0 | 8.6 |
| Relative percentage change (=Z/X*100) | 90.9 | 82.7 |

*1 USD = 29.07 Birr (2019, purchasing power parity, 1 international dollars=10.74 Birr)*

Supplementary table3.Sources of payment for surgical care

| Sources of payment | Frequency | Percentage |
| --- | --- | --- |
| Income | 97 | 53.3 |
| Sold items | 69 | 37.9 |
| Relatives | 28 | 15.3 |
| Reimbursement | 13 | 7.1 |
| Saving | 12 | 6.5 |
| Borrowing | 9 | 4.9 |
| Others | 16 | 8.8 |

Total % is more than 100 as payment is made by more than one source
